# The Neural Correlates of Autonomic Interoception in a Clinical Sample: Implications for Anxiety

**DOI:** 10.1101/2023.05.25.23290230

**Authors:** Poppy Z Grimes, Christina N Kampoureli, Charlotte L Rae, Neil A Harrison, Sarah N Garfinkel, Hugo D Critchley, Jessica A Eccles

## Abstract

Interoceptive mismatch is a perceptual discrepancy between ascending bodily signals and higher-order representation of anticipated physiological state. Inspired by predictive coding models, we present *autonomic perceptual mismatch* as a measure of this discrepancy for clinical application to brain-body interactions. Joint hypermobility is disproportionately found in individuals with anxiety disorders. Previous work has shown atypical autonomic reactivity represents a likely mediating mechanism consequent of altered connective tissue in the vasculature and nervous system.

This fMRI study investigates the neural substrates of autonomic perceptual mismatch on affective processing in the hypermobility-anxiety interaction. We compared regional brain activity during emotional face processing in participants with and without hypermobility and generalized anxiety disorder diagnosis, then tested association with perceptual mismatch.

In the brain, autonomic perceptual mismatch correlated with enhanced activation in emotion processing and autonomic control regions, notably anterior cingulate cortex. Anxious individuals exhibited increased mid-insula cortex activity in relation to perceptual mismatch. Activity was decreased within the inferior frontal gyrus, a region implicated in cognitive control. Dysautonomia mediated the link between hypermobility and anxiety.

Together, these findings support a neural basis of an autonomic perceptual mismatch model in a clinical sample. This is supported by the engagement of neural systems for emotion-cognition and interoception. This work highlights convergent aspects of neurodiversity, mental health, connective tissue disorders and brain-body interactions relevant to precision healthcare.

## 1. Introduction

The coupling of brain and body is evident in cognitive and emotive responses to visceral afferent signals. Interoception, the internal sensing of our milieu intérieur, its representation in brain, and its impact on psychological processes including bodily feelings, is proposed to be central to emotion [1–4]. Afferent interoceptive information is conveyed centrally by viscerosensory nerves that permit the reflexive and allostatic [5] autonomic nervous control of internal physiology [6]. Dysautonomia describes perturbation of adaptive autonomic control and may arise through aberrant interoceptive signalling, representation and regulation. Notably, dysautonomic symptoms are common to many mental and physical conditions.

Anxiety is commonly associated with heightened states of autonomic arousal linked to anticipatory fear and worry. The physiological signalling and perception of arousal amplify negative feelings as subjective anxiety symptoms and associated avoidant behaviours [7, 8]. Anxiety is thus linked to uncertainty regarding internal states (generalised anxiety disorder) or external situations (panic and social anxiety disorders) [9]. Anxious feelings may arise from unexplained arousal and the mismatch between anticipated/desired actual interoceptive signalling [10]. Correspondingly, discrepancies in accurately discerning interoceptive signals provide a mechanistic hypothesis for the precipitation of anxiety by dysregulated bodily states [3, 11, 12].

Interoception provides a conceptual framework for the relationship between autonomic dysfunction and anxiety [3, 9, 10, 13]. Recent studies link the expression of anxiety to attenuated interoceptive awareness (interoceptive metacognitive insight), computed as the correspondence (or mismatch) between objective measures of a person’s interoceptive sensitivity (from performance accuracy on interoceptive tasks) and their subjective perception of their own interoceptive sensitivity (rated confidence in interoceptive performance accuracy) [14]. Interoceptive mismatch has been conceptualised as the discrepancy between self-reported awareness measures of interoception (e.g. assessed through the awareness subscale of the Porges Body Awareness Questionnaire; [15]) relative to behavioural accuracy on interoceptive tests (e.g. heartbeat perception). Interoceptive mismatch is also associated with heightened anxiety symptomatology [16].

Joint hypermobility is an outward manifestation of a more general variation in the structural integrity of connective tissues, including collagen [17]. Although hypermobility is common (present in roughly 20% of the general population) some individuals develop symptoms that can affect multiple bodily systems: hypermobility is associated with chronic pain, fatigue, gastrointestinal disturbance, neurodevelopmental and neuropsychiatric conditions [18–20]. While typically underdiagnosed, the complex comorbidities of symptomatic hypermobility can go unrecognised in individuals, hindering quality-of-life [18, 21–23].

Hypermobility is highly prevalent in up to 70% of individuals clinically diagnosed with anxiety [17, 24–26]. Hypermobile individuals experience autonomic symptoms that can amplify affective anxiety [23]. Dysautonomia in hypermobility is thought to manifest consequent of reactive autonomic regulation of less elastic vascular tissues [8, 27, 28]. Imprecise feedback control of peripheral blood flow putatively results in physiological symptoms and compensatory autonomic and behavioural responses.

Despite findings of the association between hypermobility and anxiety extending to autonomic and somatic symptoms [28], a neurobiological account is yet to be elucidated. Atypical autonomic function (dysautonomia), in the interoceptive framework, may provide a mechanistic explanation.

Hypermobile individuals show higher subjective sensitivity to interoceptive sensations [27] and interoceptive accuracy influences the relationship between hypermobility and anxiety level [29]. Based on these observations, we predict the important interaction between autonomic interoception, hypermobility and anxiety will be evident in distinct brain activation patterns.

Orthostatic intolerance (OI) is the onset of autonomic symptoms upon standing, including palpitations and dizziness, linked with increased heart rate and low blood pressure [30]. OI quantifies dysautonomia [12, 13, 31] and has a 78% prevalence in hypermobility [32]. OI symptoms rise in anxious individuals [13], notably mediating the relationship between hypermobility and anxiety diagnosis [19], suggesting altered connective tissue affects autonomic function.

Functional magnetic resonance imaging (fMRI) has shown engagement of the same brain regions during anxiety states that support interoceptive representations and autonomic control in the limbic system [9]. The insula cortex is a putative mismatch computation site, shown to respond to altered physiological feedback and relay information to anterior cingulate cortex [33, 34].

Hypermobility presence increases reactivity within insula and related anxiety-associated regions [29]. Hypermobility has also been associated with increased bilateral amygdala volume [27] and insula structural differences, which correlate with increased orthostatic heart rate and interoceptive accuracy in anxious-hypermobile participants [35]. However, no brain imaging studies have examined neural mechanisms that may link hypermobility to anxiety through autonomic interoception in a clinical sample.

To investigate interoceptive mismatch in dysautonomia, we developed an interoceptive measure of autonomic perceptual mismatch (APM) – corresponding to the magnitude of mismatch in objective and subjective sensitivity to autonomic signals. This fMRI study investigates the functional neural correlates of autonomic perceptual mismatch using the hypermobility-anxiety interaction as a clinical model.

## 2. Materials and Methodology

### Participants and psychometric measures

Fifty-one participants, matched for age and gender, were recruited to the study at the Clinical Imaging Sciences Centre at the University of Sussex, Brighton, UK (**Table 1**). Of the participants, 26 had a diagnosis of generalized anxiety disorder (GAD; DSM-IV) as confirmed by a clinician using the Mini-International Neuropsychiatric Interview (MINI; [36]). The remaining 25 participants were healthy controls with no diagnosed psychiatric condition. The Brighton diagnostic criteria [37] were applied to the classification of hypermobility. Of those with a diagnosis of generalised anxiety disorder, 18 were classified as hypermobile. Six of the healthy controls were classed as hypermobile.

**Table 1.** Anxiety diagnosis (GAD) case and control demographics for hypermobility status, age and gender.

|  | % (N) | % hypermobile (N) | mean age (SD) | % female (N) |
| --- | --- | --- | --- | --- |
| Anxiety diagnosis | 51 (26) | 69 (18) | 41.4 (12.2) | 65 (17) |
| Non-anxious controls | 49 (25) | 24 (6) | 37.8 (14.4) | 44 (11) |
| total | 100 (51) | 47 (24) | 39.6 (1.9) | 55 (28) |

Participant exclusion criteria included MRI incompatibility, neurological condition and any other psychiatric condition except anxiety and depression in the anxious group. In addition to categorical classification of anxiety and/or hypermobility status, the Beck Anxiety Inventory (BAI; [38]) and the Beighton scale of joint laxity (BS; [39]) were used to quantify the degree of anxiety and hypermobility respectively. Subjective scores for orthostatic intolerance were recorded using the orthostatic subscale of the Autonomic Symptoms and Quality of Life scale (AQQoL; [40]).

Clinically anxious participants were recruited from Sussex Partnership NHS trust and via electric bulletin boards. Controls were recruited via bulletin boards at Sussex and Brighton Universities. The study procedure was ethically approved by the Brighton and Hove NRES committee (ref 12/LO/1942).

### Statistical analyses

Behavioural results were first assessed using bivariate correlations between continuous variables, autonomic perceptual mismatch and anxiety score. Subsequently, independent samples t-tests (two-tailed) were performed to investigate differences in mean autonomic perceptual mismatch scores for hypermobile and anxious participants versus controls. Psychophysiological interactions were computed using univariate interaction analyses in the General Linear Model (GLM). Variables were entered as fixed factors (categorical) or dependent variables (continuous), and perceptual mismatch was entered as the covariate. Sum-of-squares Type III method was used with intercept included in the model.

### Autonomic perceptual mismatch computed as orthostatic intolerance mismatch

Objective autonomic testing was performed using an active stand test of orthostatic intolerance, which measured heart rate (HR) change from lying down to one minute of standing. Participant heart rates were recorded using a pulse oximeter (NONIN, Nonin Medical, Minnesota, USA). Heart rates from lying to standing were recorded at baseline, peak and after one minute of standing. Changes in heart rate for each participant were calculated as absolute values and proportional to baseline for peak rate or rate after standing. Subjective scores for orthostatic intolerance were recorded based on the autonomic subsection of the AQQoLS.

Autonomic perceptual mismatch was computed as the mismatch between signs (OI stand test) and symptoms (AQQoLS) of orthostatic intolerance in the same framework as interoceptive trait prediction error [14]. This was calculated as

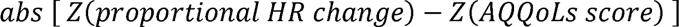

where *Z* is the standard Z-score, (*x* - *µ*)/*σ*. Absolute values were used to investigate the magnitude of error with neural activation. Final APM scores were assigned to participants as the transformed mismatch between orthostatic intolerance signs and symptoms based on predictive coding models. The results of all autonomic testing are available in Supplementary Table 1.

### Neuroimaging paradigm: Stimuli and experimental design

The in-scanner task was modified from Umeda *et al.* [41]. Emotional faces were selected and grouped into five classes (angry, afraid, disgusted, neutral, happy) from the Karolinska Directed Emotional Faces (KDEF; [42]). Faces were presented in an event-related design and were randomised and counter-balanced across runs. Attendance to faces was ensured by asking participants to determine whether they could see teeth in the faces or not. In total there were 96 trials per participant; there were 15 trials per class of emotional face and 21 fixation cross trials used as the implicit baseline which served as the control condition. Each stochastically ordered trial was 4 seconds long, during which the face remained on screen, and the participant was expected to respond. The fixation cross duration was also 4 seconds in duration. A total run lasted 384s (6m 24s) and each participant underwent two runs of the emotional faces task.

### Image acquisition

Neuroimaging took place using a 1.5 Tesla Siemens Avanto Scanner with a 32-channel head coil (Siemens Medical Solutions, Erlangen, Germany). T1 structural scans were first acquired for each participant using a magnetisation-prepared rapid gradient-echo acquisition (repetition time TR = 2.73s per volume, echo time TE = 3.57msec, inversion time = 1000ms, flip angle = 7°). T2*-weighted whole-brain functional scans were taken using a single-shot 2D gradient echo-planar imaging (EPI) sequence. For the functional scans, voxel sizes were 3×3×3mm, repetition time TR = 2.52s per volume, echo time TE = 43ms. 34 axial slices of 3mm thickness and 0.6mm interslice gap were taken. Slices were tilted at a 30° flip angle from the intercommissural plane to minimise signal artefacts.

### Pre-processing

Imaging data were processed in SPM12 (http://www.fil.ion.ucl.ac.uk/spm) on MATLAB R2021b (v9.9.0 Mathworks Inc.). Echo planar images were realigned to the mean image for motion correction, scanner drift and variation. Slice-time correction to slice 6 (which aligned with amygdala) and was performed for all volumes to remove artefacts. The first two volumes were discarded for scanner equilibration. T2*-functional scans were co-registered with T1-structural scans for each participant. Images were normalised to the MNI-152 (Montreal Neurologic Institute) brain space. The data were smoothed spatially with a Gaussian kernel of 8 mm full-width half-maximum.

### Univariate neuroimaging analysis

First- and second-level designs were implemented and analysed under the general linear model (GLM) in SPM12 on the whole brain. First-level modelling included a regressor for each of the five emotional face conditions and the implicit baseline (control condition) of each individual. Participant motion from image realignment (3 translations and 3 rotations) was included as six regressors of no interest for each participant at the first level. Statistical comparison of voxel-wise parameters was conducted for first-level contrasts within subjects (emotional face>baseline), and these contrasts for the five emotions were entered into the second level where a full-factorial design (2×2×5) was used for component interaction analysis.

The first factor had two levels (hypermobile, non-hypermobile), the second factor had two levels (anxious, non-anxious) and the third factor had five levels (angry, afraid, happy, neutral, sad). Regressors were entered for age and gender. In this second-level model, autonomic perceptual mismatch was added as a covariate to investigate the correlations and interactions of autonomic perceptual mismatch on hypermobility and anxiety. All covariates were mean-centred at zero.

Statistical comparison of voxel-wise parameters was conducted for second-level contrasts between subjects (anxious>non-anxious), (hypermobile>non-hypermobile). A family-wise error cluster correction (FWEc) was performed on the whole brain at *P<*0.05 to correct for multiple comparisons. This reduced the likelihood of type-I errors, thus minimising false positives. Brain activations were interpreted as clusters that produced a significant change in BOLD response to the emotional faces fMRI task (relative to the baseline of fixation cross).

Clusters were anatomically defined using the SPM Anatomy toolbox v3.0 [43]. Where labelling was not available in the anatomy toolbox, the Harvard-Oxford anatomical atlas was used by overlaying the relevant t-contrasts onto the ‘ch2better’ template on MRIcron v1.0.20190902 [44].

For each participant, the eigenvariates were extracted (weighted mean of Blood-Oxygen-Level-Dependency (BOLD) time series) from the peaks of activation. Eigenvariates were averaged over the five conditions for each participant for each t-contrast of interest and used to generate scatter plots against autonomic perceptual mismatch.

### Mediation

The interaction between hypermobility, autonomic perceptual mismatch and anxiety was investigated further as a secondary analysis using a Baron and Kenny [45] mediation analysis. Mediation testing conducts three regression correlations between pairs of variables. If when controlling for one factor (the potential mediator), the correlation between the remaining factors is reduced, then the controlled factor is said to partially mediate their relationship. Based on previous findings that autonomic dysfunction mediates the anxiety-laxity link [19], we entered APM as our model mediator. Consistent with development chronology, hypermobility (status and score) was employed as the independent variable, appearing in early childhood [26, 46]. Anxiety manifests later, thus being entered as the dependent or outcome variable.

## 3. Results

### Autonomic testing for orthostatic intolerance mismatch

Both signs (Fig. 1A) and symptom scores (Fig. 1B) of orthostatic intolerance were higher in anxious-hypermobile participants indicating a possible ‘mismatch error’ of autonomic dysfunction.

**Figure 1.**
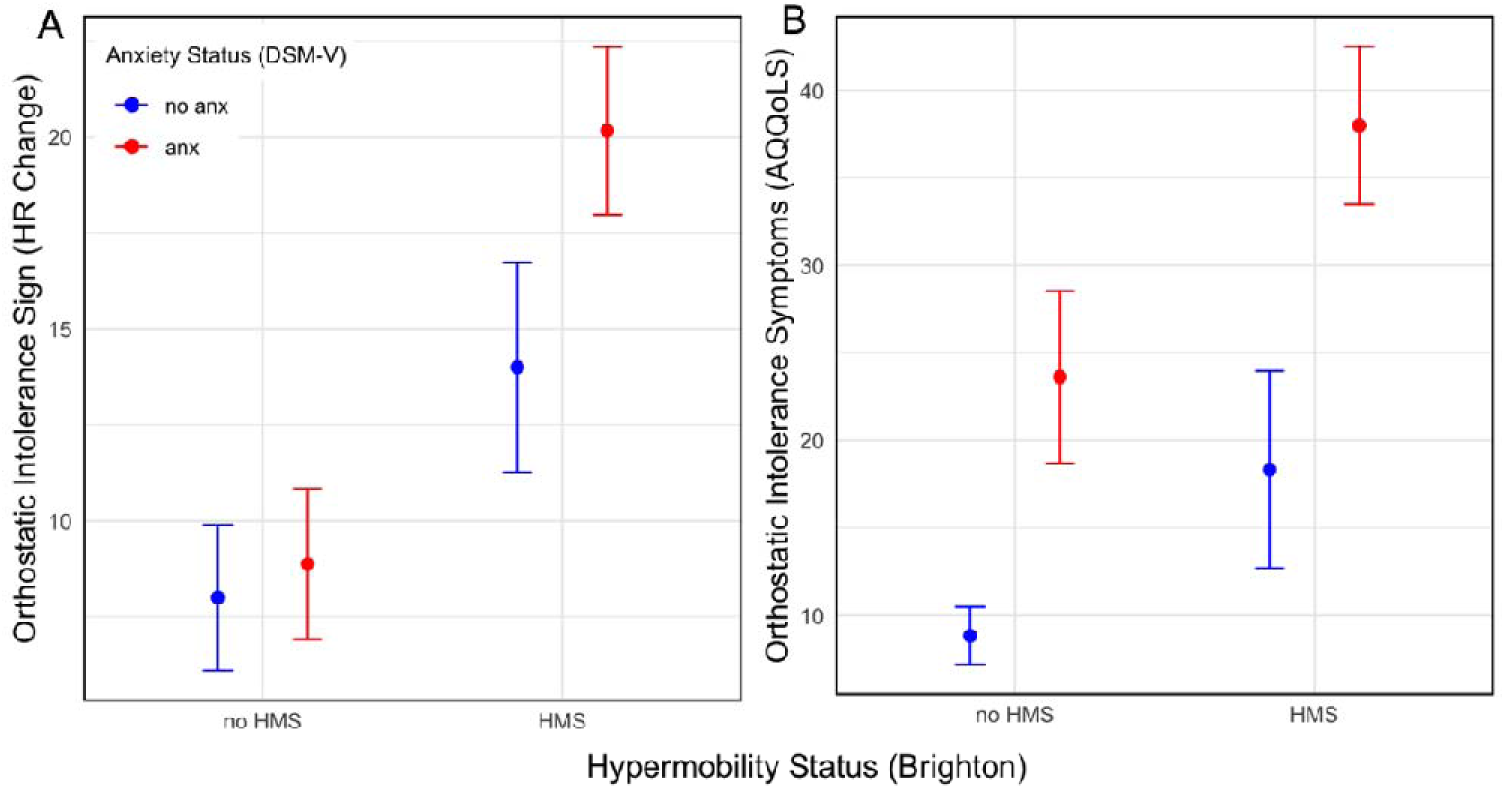
Orthostatic Intolerance (OI) in Hypermobility and Anxiety. Objective OI sign measured as the absolute heart rate change after 1 min sitting to standing (A). Subjective OI symptom score from the AQQoLS (B). Error bars represent ±1SE.

### Phenotypic correlations of autonomic perceptual mismatch

Anxiety score was positively correlated with autonomic perceptual mismatch across all participants (R_pcc_=0.425, P=0.002, Fig. 2A). When subgrouping by clinical anxiety status, the positive correlation of autonomic perceptual mismatch with anxiety score was significantly higher in those with anxiety (independent samples t-test; t=-2.964, p= 0.005, SED=0.149, Fig. 2B). Similarly, the hypermobile group showed a significantly stronger positive correlation between anxiety score and autonomic perceptual mismatch (independent samples t-test; t=-2.13, p=0.038, SED=0.155, Fig. 2C).

**Figure 2.**
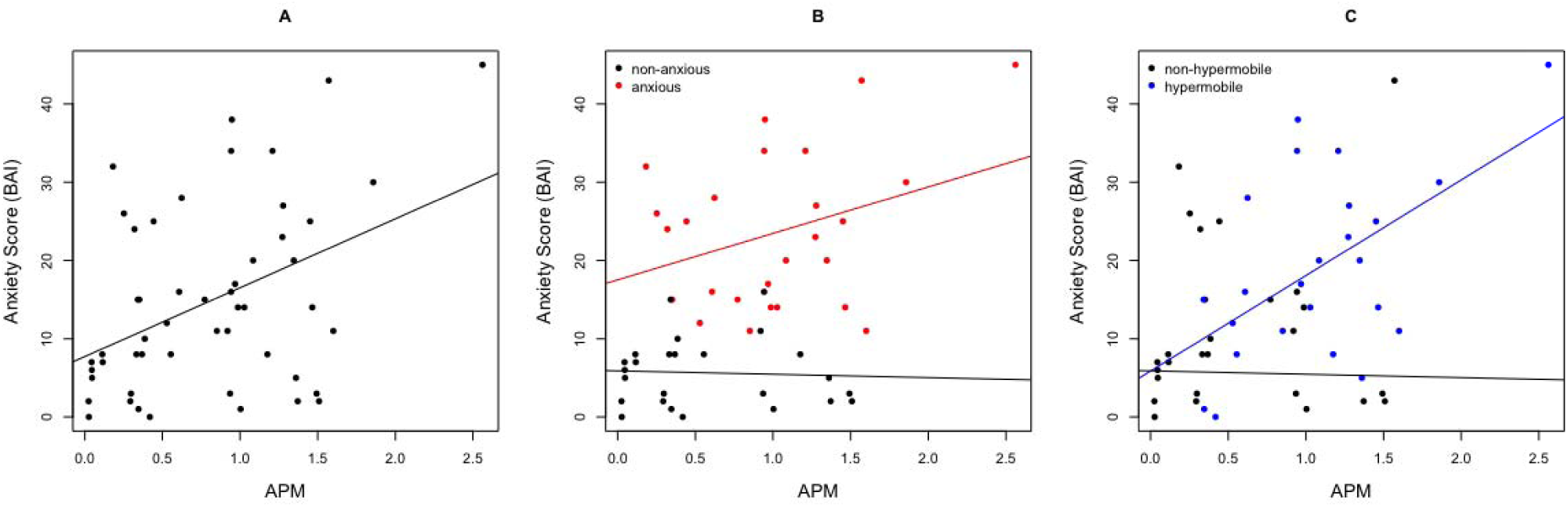
Autonomic perceptual mismatch in the hypermobility-anxiety association. Autonomic perceptual mismatch correlates with anxiety score (A). Anxiety score is higher with increased autonomic perceptual mismatch in clinically anxious (B) and hypermobile (C) individuals.

GLM univariate analysis demonstrated a significant interaction between hypermobility and anxiety on autonomic perceptual mismatch (Fig. 3, F=6.04, p=0.002). A significant interaction between hypermobility and autonomic perceptual mismatch was found on anxiety status (F=7.66, p=0.008). This interaction effect was also found on anxiety score (F=4.20, p=0.046) with a main effect of autonomic perceptual mismatch (F=5.70, p=0.021). There was no significant interaction between anxiety status and autonomic perceptual mismatch on hypermobility.

**Figure 3.**
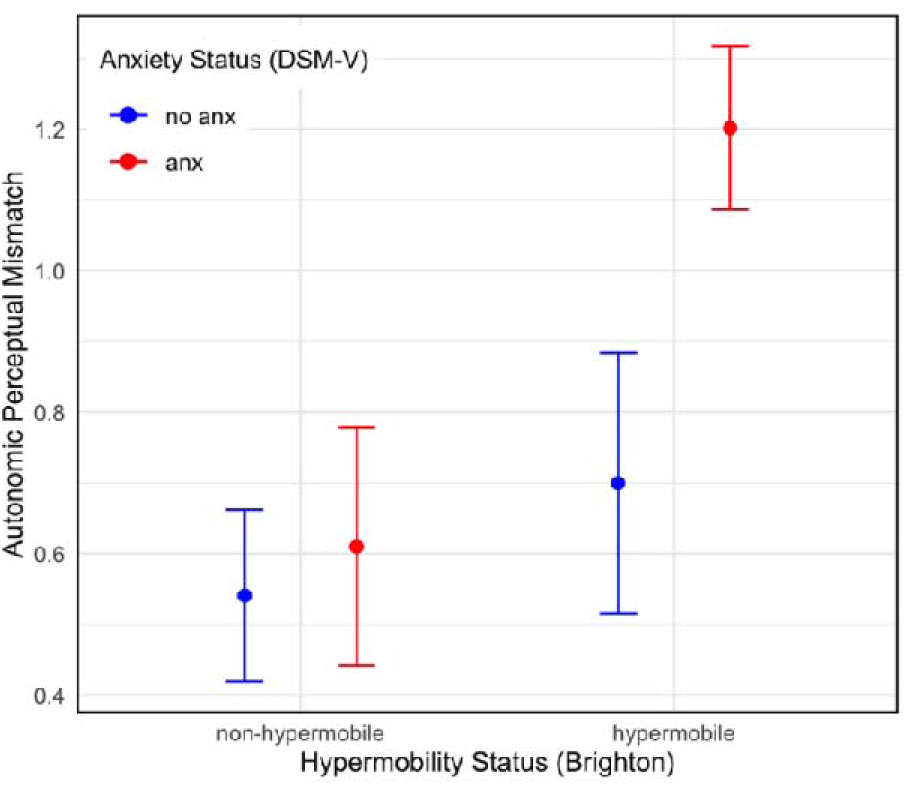
The hypermobility-anxiety interaction on autonomic perceptual mismatch. Participants classed as anxious-hypermobile have significantly higher APM scores. Bars represent +/- 1 SEM.

The APM score for one participant fell above the 95% percentile (APM = 2.56, Supplementary Table 2). Although still representable of an extreme case of perceptual mismatch in anxiety, we excluded the results of this individual from subsequent statistical neuroimaging analyses. The significant interaction in Fig. 3 between anxiety and hypermobility on APM remained after exclusion of this participant from the data.

### Univariate functional neuroimaging results

#### Neural correlations with autonomic perceptual mismatch

The first t-contrasts modelled the BOLD response to emotional faces to identify significant clusters of activation that varied with autonomic perceptual mismatch when added as a covariate to the second-level model. Autonomic perceptual mismatch correlated with response to emotional faces in the inferior frontal gyrus (pars triangularis and pars opercularis) and middle insular cortex. Full results of all imaged activations are available in Supplementary Table 2.

### Interaction with anxiety

The interaction of anxiety and autonomic perceptual mismatch produced a positive response (regions showing increased activation with APM in anxious participants) within the right anterior cingulate gyrus (Fig. 4A) and left middle insular cortex (Fig. 4B). Autonomic perceptual mismatch produced a negative response in the right inferior frontal gyrus (Fig. 4C) and anterior mid-cingulate cortex (Fig. 4D) in anxious participants. Weighted mean BOLD estimates for the anterior cingulate gyrus and mid-insula were positively correlated with APM; inferior frontal gyrus and mid-cingulate estimates were negatively correlated with APM in anxiety (Fig. 4E).

**Figure 4.**
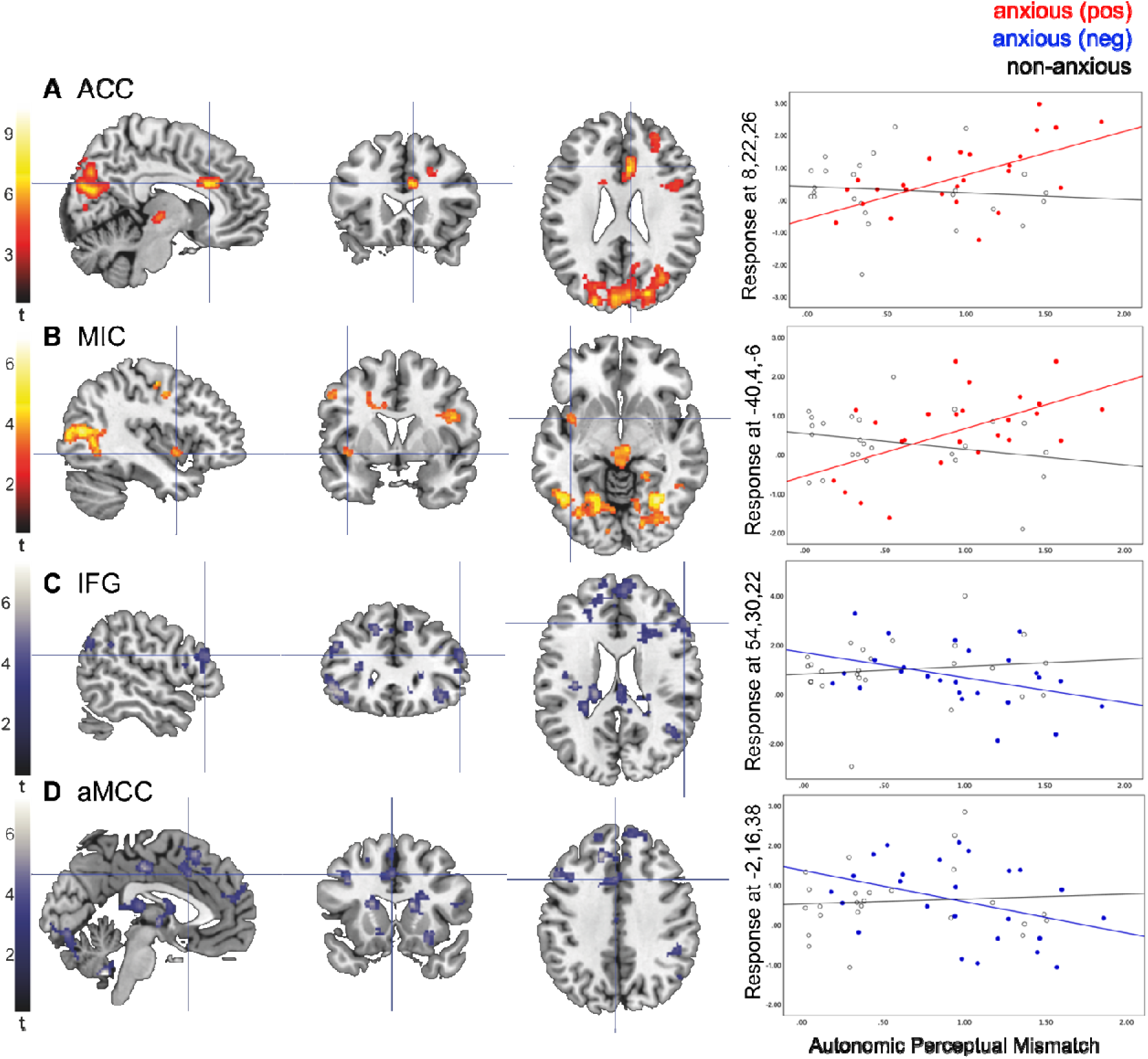
T-contrast estimates of the interaction between anxiety and autonomic perceptual mismatch. Emotional face stimuli increased activation in the anterior cingulate (A) and mid-insula (B). Activation was decreased in the IFG (C) and mid-cingulate (D). Colour bars represent peak-level t-statistics at FWEc P<0.05. Crosshairs represent MNI co-ordinates of cluster peak activation. Scatter plots show the interaction between APM and anxiety at mean peak-clusters of activation from extracted eigenvariates for positive and negative responses in anxious individuals vs. controls (E).

### Interaction with hypermobility

The interaction of hypermobility and autonomic perceptual mismatch produced a positive response (regions showing increased activation with APM in hypermobile participants) in the left mid cingulate gyrus (Fig. 5A). Autonomic perceptual mismatch produced a negative response in the left inferior frontal gyrus in hypermobile participants (Fig. 5B). Weighted mean BOLD estimate for the anterior cingulate gyrus was positively correlated with autonomic perceptual mismatch and inferior frontal gyrus estimates was negatively correlated in hypermobility (Fig. 5C).

**Figure 5.**
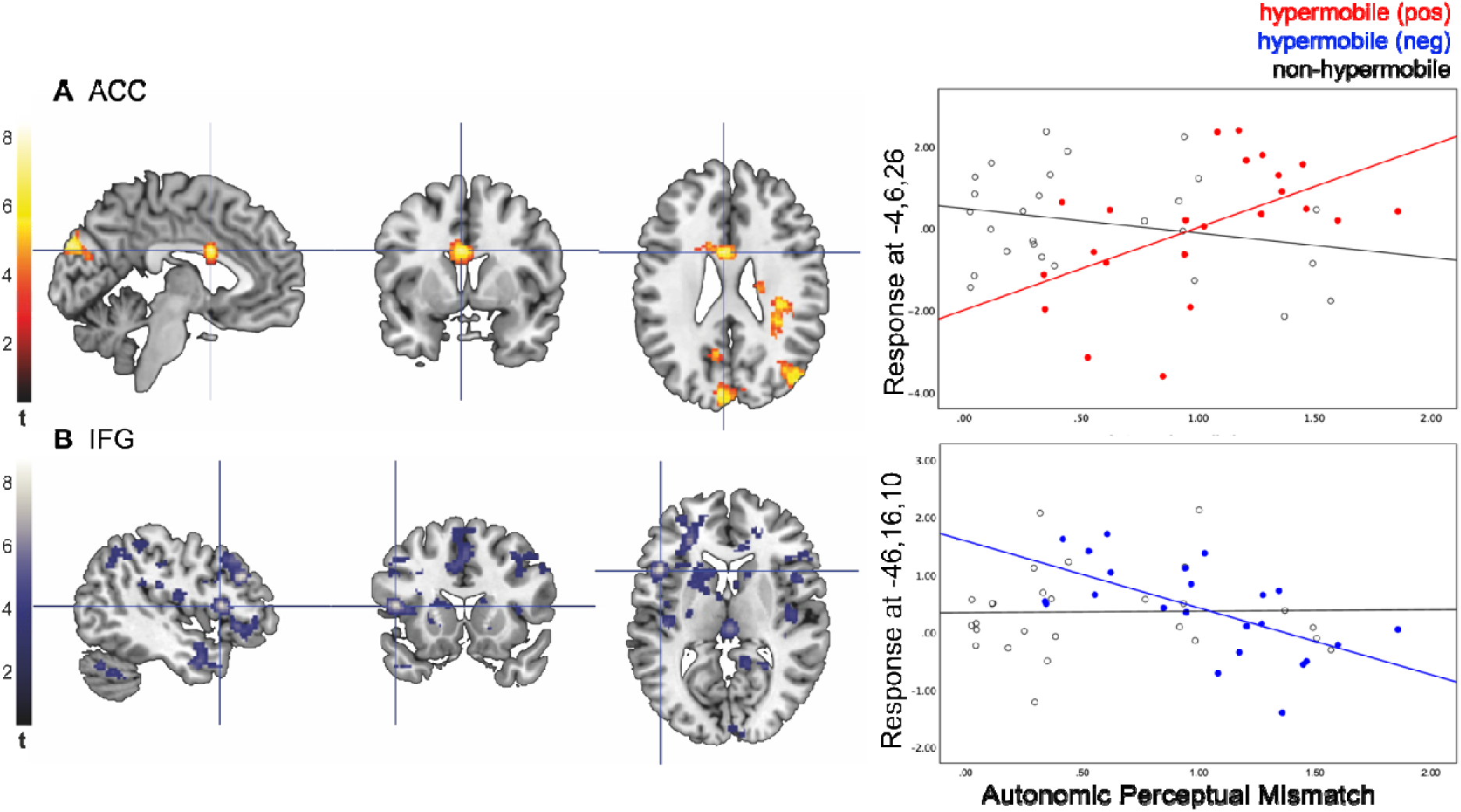
T-contrast estimates of the interaction between hypermobility and autonomic perceptual mismatch. Emotional face stimuli induced an increased response in the **anterior cingulate** (A) and reduced response in the IFG (B). Colour bars represent peak-level t-statistics at FWEc P<0.05. Crosshairs represent MNI co-ordinates of cluster peak activation. Scatter plots show the interaction between APM and hypermobility at mean peak-clusters of activation from extracted eigenvariates for positive and negative responses in anxious individuals vs. controls (C).

### Mediation

Upon mediation testing, we found autonomic perceptual mismatch to partially mediate the relationship between hypermobility and anxiety status (Fig. 6A; categorical variables) when controlled for. Autonomic perceptual mismatch fully mediated the relationship between hypermobility and anxiety scores (Fig. 6B; continuous variables) when controlled for.

**Figure 6.**
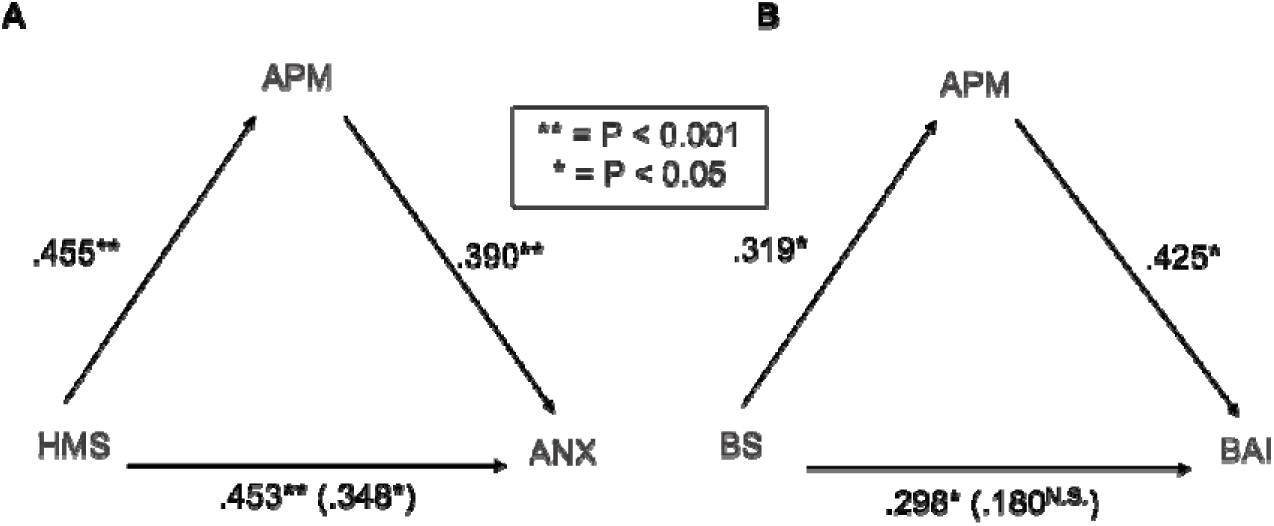
Baron and Kenny mediation. Partial mediation of APM on effect of hypermobility status on anxiety status (A). Full mediation of APM on effect of Beighton score on Beck’s anxiety score (B). Reported correlations are standardised beta coefficients.

## 4. Discussion

This study has shown that autonomic perceptual mismatch – a measure of autonomic dysfunction inspired by predictive coding models – implicates functional brain activity and is positively associated with anxiety. In a clinical sample, we provide evidence that dysautonomia may be a mechanism by which hypermobility evokes anxiety through a discrete set of brain regions that support interoceptive representation and autonomic control.

Our findings are consistent with prior research which showed differences in brain activity and a mediating effect of objective interoception on anxiety, in a non-clinical hypermobile group [29]. However, this previous study did not consider the mismatch between objective and subjective awareness.

### Altered emotion-autonomic processing in anterior cingulate cortex

Increased anterior cingulate cortex (ACC) activation was observed in both anxious and hypermobile participants with autonomic perceptual mismatch. ACC is implicated in emotion processing, particularly negative emotions like anxiety, and interoceptive accuracy [3, 47–49]. Imaging studies demonstrate high ACC activity in people with high trait anxiety and involvement in the physiological response to stress as a locus of autonomic regulation [50–54]. Our findings support the significant role of ACC in emotion-autonomic regulation, contributing to the development and persistence of anxiety disorders.

### Mid-insula as a locus linking interoception and anxiety

Interestingly, we found a positive response to emotional faces in the middle insula cortex in the interaction of APM with anxiety. Mid-insula is often overlooked in interoception research, as the focus is usually on anterior insula as the mismatch comparator [10, 33, 55, 56]. However, some hypotheses suggest mid-insula to implicate polymodal integration of emotion-interoception processing, with somatosensory and bodily afferent information projecting to the anterior insula to inform emotional experience and intensity [57–61]. Functional connections also exist to cingulate and frontal cortices, where autonomic control pertaining to interoceptive information is elicited and feedback integrated [55, 62–64].

Mid-insula is commonly activated in interoceptive neural activations across psychiatric conditions [65]. Its activity correlates with interoceptive accuracy and anxiety scores, indicating a role in anxiety-related interoception [34]. The insula is functionally graded, with the mid lobe as an intermediary structure [66]. Interoceptive responses to bodily awareness and organ distension follow this graded response [67].

Paulus and Khalsa [55] propose re-evaluating the mid-insula’s role in integrating autonomic-interoceptive information. Our observed activity patterns suggest potential alterations of this integration, which could result in mismatched models in the brain and symptom anxiety, corresponding to interoceptive predictive coding. The mid-insula’s connections to cingulate and frontal regions may contribute to this effect.

### Reduced frontal control may predispose anxious symptoms

Our study revealed reduced response in a distinct cluster in the anterior mid-cingulate cortex (aMCC) in the interaction of APM with anxiety. aMCC is linked to cognitive function and error detection, co-activating and functionally connected to insula cortex [1, 50, 62, 68– 70]. Important for pain perception [71–74], attenuation of aMCC activity demonstrated reduced awareness of error detection [75]. aMCC is also a key locus of the Salience Network for internal-external stimulus recognition [33, 76]. In line with this work, our observation suggests that reduced activity of the mid-cingulate may be a locus of autonomic perceptual mismatch. Connectivity and co-activation to insula may be a path to the onset of subsequent anxious symptoms.

We observed reduced activation of bilateral inferior frontal gyri (IFG) with autonomic perceptual mismatch in both hypermobile and anxious individuals. IFG is associated with a variety of cognitive functions such as emotion regulation and response inhibition [77, 78]. IFG is functionally connected to the insula, playing a role in viscero-motor-autonomic functioning [79]. Thus, perceptual mismatch may implicate these functional networks, resulting in altered autonomic function and related cognitive processes. The accumulation of interoceptive errors may contribute to decision-making errors at a higher level, potentially exacerbating anxiety. However, further research is needed to investigate the mechanisms underlying emotion-autonomic processing in networks involving IFG and insular cortex.

### Dysautonomia as a mediator in brain-body interactions

We demonstrate that autonomic perceptual mismatch mediates the effect of hypermobility on anxiety. Our study adds to existing evidence that interoceptive accuracy [29] and orthostatic symptoms [19] mediate this association. We have employed the interoceptive mismatch framework [16], to elucidate the underlying mechanism of this relationship. Our findings support the hypothesis that dysautonomia in hypermobile individuals, resulting from altered vascular collagen [8], may explain the observed overlap between hypermobility and anxiety.

### Clinical relevance to the anxiety-hypermobility link

The link between connective tissue disorders and psychiatric conditions requires a holistic approach to understanding brain-body interactions. The ‘Neuroconnective phenotype’ [25, 80] is a clinical recognition of the hypermobility-anxiety link that combines somatic and sensory symptoms, behaviour, psychiatry and neurodevelopmental conditions [20, 81]. Autonomic symptoms are higher in neurodevelopmental conditions [19, 81, 82] and autism is overrepresented by up to 52% in hypermobility [83].

Our work utilising the Neuroconnective model suggests that somatic symptoms may be linked to differences in sensory processing, which is common to autism and hypermobility, but needs further empirical research. These findings highlight the need for personalised screening for various conditions in those displaying autonomic dysfunction in hypermobility.

One limitation is that we did not consider the influence of psychotropic medication on our results which may impact on autonomic symptoms to varying degrees [8]. Nonetheless, Eccles [8] demonstrated that robust differences in autonomic function of hypermobile individuals exist regardless of medication status. Exploring medication effect on brain function in the context of hypermobility and autonomic dysfunction may implicate personalised treatments and phenotyping that account for individual responses to psychotropic medication.

### Conclusions

Our study demonstrates how autonomic dysfunction affects neural activity in emotion-processing and cognitive control regions in hypermobility and anxiety. These findings offer a mechanistic explanation by which imprecise autonomic interoceptive signals from bodily afferents manifest as anxiety through autonomic mediation. The involvement of mid-insula in polymodal error weighting provides novel evidence for interoceptive predictive coding models. These findings have important implications for brain-body interactions in connective tissue disorders contributing to anxiety symptomatology.

## Data Availability

All data in the study can be found in the supplementary materials or are available upon reasonable request to the authors.

## Author Contributions

PZG, JAE and HDC conceptualised the project and methodology. PZG was responsible for data analysis, manuscript writing and visualisation. JAE was responsible for the initial recruitment and fMRI data acquisition. CLR, NAH and HDC contributed to the methodology. JAE acquired funding for the project. All authors contributed to manuscript editing and reviewing.

## Declaration of interest

All authors declare no conflict of interest.

## Sources of Funding

Funding for this project came via a fellowship to JAE (MRC MR/K002643/1). JAE was also supported by MQ Transforming Mental Health and Versus Arthritis (MQF 17/19).

## Notes

### Competing Interest Statement

The authors have declared no competing interest.

### Author Declarations

Brighton and Hove National Research Ethics Service (NREC) committee gave ethical approval for this work (ref 12/LO/1942).

### Summary of Updates

Manuscript abstract and introduction revised

