## Supplementary Tables for "The Neural Correlates of Autonomic Interoception in a Clinical Sample: Implications for Anxiety"

| HMS | ANX | OI symptom score | Baseline HR | Initial HR | 1 min HR | ABS 1 min change HR | Proportional HR change | z score OI sign | z score OI symptom | Raw APM | Trans APM |
| --- | --- | --- | --- | --- | --- | --- | --- | --- | --- | --- | --- |
| 1 | 2 | 20 | 64 | 80 | 77 | 13 | 0.2 | 0.04474 | -0.13737 | 0.18 | 0.18 |
| 2 | 1 | 2 | 64 | 137 | 66 | 2 | 0.03 | -1.12462 | -1.09999 | -0.02 | 0.02 |
| 2 | 1 | 17 | 70 | 97 | 71 | 1 | 0.01 | -1.24003 | -0.2978 | -0.94 | 0.94 |
| 2 | 2 | 22 | 43 | 63 | 63 | 20 | 0.47 | 1.82722 | -0.03041 | 1.86 | 1.86 |
| 2 | 2 | 30 | 51 | 76 | 76 | 25 | 0.49 | 1.99785 | 0.39742 | 1.6 | 1.6 |
| 1 | 1 | 12 | 47 | 54 | 55 | 8 | 0.17 | -0.17918 | -0.5652 | 0.39 | 0.39 |
| 1 | 1 | 2 | 71 | 73 | 73 | 2 | 0.03 | -1.14558 | -1.09999 | -0.05 | 0.05 |
| 2 | 1 | 13 | 69 | 112 | 74 | 5 | 0.07 | -0.84422 | -0.51172 | -0.33 | 0.33 |
| 2 | 2 | 33 | 61 | 92 | 91 | 30 | 0.49 | 2.00878 | 0.55786 | 1.45 | 1.45 |
| 1 | 2 | 23 | 89 | 51 | 101 | 12 | 0.13 | -0.4199 | 0.02307 | -0.44 | 0.44 |
| 2 | 2 | 74 | 55 | 75 | 83 | 28 | 0.51 | 2.1264 | 2.7505 | -0.62 | 0.62 |
| 1 | 1 | 10 | 74 | 95 | 80 | 6 | 0.08 | -0.78559 | -0.67216 | -0.11 | 0.11 |
| 1 | 1 | 7 | 74 | 95 | 80 | 6 | 0.08 | -0.78559 | -0.83259 | 0.05 | 0.05 |
| 2 | 2 | 27 | 112 | 121 | 121 | 9 | 0.08 | -0.79051 | 0.23699 | -1.03 | 1.03 |
| 1 | 1 | 6 | 77 | 85 | 99 | 22 | 0.29 | 0.60664 | -0.88607 | 1.49 | 1.49 |
| 1 | 1 | 1 | 63 | 80 | 52 | -11 | -0.17 | -2.52515 | -1.15347 | -1.37 | 1.37 |
| 2 | 2 | 25 | 64 | 96 | 88 | 24 | 0.38 | 1.2141 | 0.13003 | 1.08 | 1.08 |
| 2 | 2 | 36 | 90 | 113 | 135 | 45 | 0.5 | 2.06455 | 0.7183 | 1.35 | 1.35 |
| 1 | 2 | 6 | 68 | 69 | 69 | 1 | 0.01 | -1.23718 | -0.88607 | -0.35 | 0.35 |
| 1 | 1 | 11 | 64 | 83 | 68 | 4 | 0.06 | -0.91201 | -0.61868 | -0.29 | 0.29 |
| 2 | 1 | 10 | 79 | 121 | 91 | 12 | 0.15 | -0.30378 | -0.67216 | 0.37 | 0.37 |
| 1 | 2 | 66 | 62 | 87 | 82 | 20 | 0.32 | 0.85747 | 2.32267 | -1.47 | 1.47 |
| 1 | 1 | 9 | 64 | 95 | 70 | 6 | 0.09 | -0.6994 | -0.72564 | 0.03 | 0.03 |
| 1 | 1 | 0 | 60 | 79 | 70 | 10 | 0.17 | -0.2033 | -1.20695 | 1 | 1 |
| 1 | 1 | 4 | 68 | 82 | 71 | 3 | 0.04 | -1.03707 | -0.99303 | -0.04 | 0.04 |
| 2 | 2 | 61 | 81 | 112 | 107 | 26 | 0.32 | 0.84663 | 2.05527 | -1.21 | 1.21 |
| 2 | 1 | 25 | 79 | 112 | 100 | 21 | 0.27 | 0.47131 | 0.13003 | 0.34 | 0.34 |
| 2 | 2 | 12 | 65 | 76 | 65 | 0 | 0 | -1.33723 | -0.5652 | -0.77 | 0.77 |
| 2 | 1 | 42 | 68 | 88 | 80 | 12 | 0.18 | -0.1366 | 1.03917 | -1.18 | 1.18 |
| 1 | 1 | 5 | 50 | 63 | 56 | 6 | 0.12 | -0.5208 | -0.93955 | 0.42 | 0.42 |
| 1 | 2 | 44 | 82 | 97 | 93 | 11 | 0.13 | -0.42456 | 1.14613 | -1.57 | 1.57 |
| 2 | 2 | 18 | 64 | 92 | 80 | 16 | 0.25 | 0.36366 | -0.24433 | 0.61 | 0.61 |
| 2 | 2 | 51 | 57 | 92 | 73 | 16 | 0.28 | 0.57254 | 1.52048 | -0.95 | 0.95 |
| 1 | 1 | 27 | 84 | 112 | 102 | 18 | 0.21 | 0.12068 | 0.23699 | -0.12 | 0.12 |
| 2 | 2 | 55 | 80 | 94 | 86 | 6 | 0.07 | -0.82696 | 1.7344 | -2.56 | 2.56 |
| 2 | 2 | 42 | 74 | 98 | 86 | 12 | 0.16 | -0.23395 | 1.03917 | -1.27 | 1.27 |
| 2 | 1 | 12 | 67 | 96 | 88 | 21 | 0.31 | 0.79523 | -0.5652 | 1.36 | 1.36 |
| 2 | 1 | 19 | 68 | 99 | 85 | 17 | 0.25 | 0.36366 | -0.19085 | 0.55 | 0.55 |
| 1 | 1 | 4 | 53 | 78 | 58 | 5 | 0.09 | -0.69538 | -0.99303 | 0.3 | 0.3 |
| 2 | 1 | 7 | 56 | 89 | 63 | 7 | 0.13 | -0.48678 | -0.83259 | 0.35 | 0.35 |
| 2 | 2 | 25 | 84 | 112 | 99 | 15 | 0.18 | -0.12231 | 0.13003 | -0.25 | 0.25 |
| 2 | 1 | 22 | 55 | 91 | 73 | 18 | 0.33 | 0.88939 | -0.03041 | 0.92 | 0.92 |
| 1 | 1 | 8 | 79 | 111 | 103 | 24 | 0.3 | 0.72967 | -0.77912 | 1.51 | 1.51 |
| 2 | 1 | 3 | 61 | 85 | 72 | 11 | 0.18 | -0.11036 | -1.04651 | 0.94 | 0.94 |
| 2 | 2 | 12 | 68 | 90 | 81 | 13 | 0.19 | -0.03655 | -0.5652 | 0.53 | 0.53 |
| 1 | 2 | 17 | 61 | 107 | 79 | 18 | 0.3 | 0.67038 | -0.2978 | 0.97 | 0.97 |
| 1 | 2 | 15 | 92 | 112 | 100 | 8 | 0.09 | -0.72491 | -0.40476 | -0.32 | 0.32 |
| 1 | 2 | 44 | 50 | 78 | 61 | 11 | 0.22 | 0.15955 | 1.14613 | -0.99 | 0.99 |
| 2 | 2 | 61 | 74 | 112 | 101 | 27 | 0.36 | 1.11205 | 2.05527 | -0.94 | 0.94 |
| 2 | 2 | 38 | 83 | 94 | 94 | 11 | 0.13 | -0.45277 | 0.82525 | -1.28 | 1.28 |
| 1 | 2 | 16 | 64 | 117 | 81 | 17 | 0.27 | 0.49973 | -0.35128 | 0.85 | 0.85 |

**Supplementary Table 1 | Results of autonomic testing for autonomic perceptual mismatch scores.** 1 = non-hypermobile/anxious, 2 = hypermobile/anxious. OI symptom score was recorded using the AQQoLS subjective scale. Proportional HR change $= (ABS 1 min change/baseline)$. Raw APM = $Z(OI sign - OI symptom)$. Trans APM = $\surd{(\text{raw APM})}^{2}$

| Contrast | Response | k (extent threshold) | Location | K (no. of voxels) | T | Z | x | y | z |
| --- | --- | --- | --- | --- | --- | --- | --- | --- | --- |
| Main effect of APM | positive | 139 | lateral occipital cortex | 5562 | 8.56 | Inf | 36 | -72 | 38 |
|  |  |  | precuneous cortex | 843 | 8.46 | Inf | -14 | 8 | 26 |
|  |  |  | middle temporal gyrus | 469 | 8.12 | 7.6 | -52 | -62 | 2 |
|  |  |  | inferior lateral occipital cortex | 1246 | 7.11 | 6.75 | 32 | -84 | 0 |
|  |  |  | right thalamus | 251 | 6.23 | 5.98 | 10 | -16 | 4 |
|  |  |  | middle temporal gyrus | 139 | 5.66 | 5.47 | 50 | -52 | 0 |
|  |  |  | inferior frontal gyrus (pars triangularis) | 194 | 5.28 | 5.12 | -48 | 32 | 10 |
|  |  |  | angular gyrus right | 162 | 5.12 | 4.98 | 44 | -58 | 46 |
| Main effect of APM | negative | 142 | planum temporale | 1200 | 8.78 | Inf | -42 | -42 | 16 |
|  |  |  | right putamen | 1781 | 8.3 | 7.74 | 22 | 2 | 0 |
|  |  |  | brainstem | 537 | 7.96 | 7.47 | 10 | -44 | -34 |
|  |  |  | anterior supramarginal gyrus | 559 | 7.32 | 6.93 | 44 | -30 | 40 |
|  |  |  | inferior temporal gyrus | 659 | 7.02 | 6.67 | 44 | -18 | -20 |
|  |  |  | middle frontal gyrus | 176 | 6.91 | 6.57 | -44 | 28 | 30 |
|  |  |  | precentral gyrus | 201 | 6.85 | 6.52 | 22 | -12 | 66 |
|  |  |  | precentral gyrus | 747 | 6.4 | 6.13 | 62 | 10 | 24 |
|  |  |  | precentral gyrus | 337 | 6.06 | 5.83 | -6 | -26 | 64 |
|  |  |  | middle cingulate gyrus | 365 | 5.86 | 5.65 | -2 | -4 | 32 |
|  |  |  | left caudate | 385 | 6.02 | 5.79 | -14 | 14 | 12 |
|  |  |  | mid insular cortex | 142 | 5.46 | 5.28 | -38 | 2 | -16 |
|  |  |  | inferior frontal gyrus (pars opercularis) | 1079 | 5.68 | 5.49 | -50 | 16 | 10 |
|  |  |  | right amygdala | 266 | 4.73 | 4.61 | 16 | -8 | -18 |
|  |  |  | intracalcarine cortex | 862 | 5.77 | 5.57 | 4 | -88 | 0 |
|  |  |  | posterior cingulate gyrus | 283 | 5.73 | 5.53 | -14 | -42 | 36 |
| Interaction APM * HMS | positive | 121 | lateral occipital cortex | 377 | 8.15 | 7.62 | 40 | -62 | 8 |
|  |  |  | middle temporal gyrus | 484 | 7.45 | 7.04 | -52 | -62 | 2 |
|  |  |  | anterior cingulate gyrus | 210 | 7.44 | 7.03 | -4 | 6 | 26 |
|  |  |  | cuneal cortex | 337 | 7.42 | 7.01 | -6 | -88 | 28 |
|  |  |  | precuneous cortex | 170 | 6.58 | 6.29 | -22 | -58 | 16 |
|  |  |  | occipital fusiform gyrus | 133 | 6.46 | 6.18 | -28 | -62 | -6 |
|  |  |  | parietal operculum cortex | 312 | 6.26 | 6.01 | 36 | -28 | 26 |
|  |  |  | lingual gyrus | 121 | 6.19 | 5.95 | 22 | -40 | -8 |
|  |  |  | left thalamus | 134 | 4.17 | 4.09 | -14 | -24 | 12 |
|  |  |  | precentral gyrus | 137 | 5.82 | 5.62 | -28 | -26 | 48 |
| Interaction APM * HMS | negative | 126 | paracingulate | 585 | 8.91 | Inf | 8 | 24 | 42 |
|  |  |  | lateral occipital cortex | 2459 | 8.31 | 7.75 | -32 | -62 | 44 |
|  |  |  | right cerebellum | 829 | 8.26 | 7.71 | 26 | -88 | -38 |
|  |  |  | middle frontal gyrus | 601 | 8.14 | 7.61 | -48 | 30 | 28 |
|  |  |  | inferior frontal gyrus (pars opercularis) | 2877 | 7.99 | 7.49 | -46 | 16 | 10 |
|  |  |  | lateral occipital cortex | 546 | 7.44 | 7.03 | 24 | -56 | 34 |
|  |  |  | right putamen | 202 | 6.63 | 6.33 | 24 | 0 | -2 |
|  |  |  | temporal pole | 278 | 6.58 | 6.29 | -44 | 4 | -28 |
|  |  |  | posterior cingulate gyrus | 2473 | 6.51 | 6.22 | 14 | -46 | 6 |
|  |  |  | precentral gyrus | 560 | 6.46 | 6.18 | -16 | -26 | 42 |
|  |  |  | frontal pole | 134 | 6.21 | 5.96 | 30 | 50 | -8 |
|  |  |  | precentral gyrus | 141 | 6.18 | 5.93 | 58 | -6 | 42 |
|  |  |  | left cerebellum | 333 | 5.69 | 5.5 | -22 | -74 | -42 |
|  |  |  | central opercular cortex | 136 | 5.6 | 5.42 | 46 | -14 | 12 |
|  |  |  | lingual gyrus | 416 | 5.47 | 5.3 | 6 | -84 | -2 |
|  |  |  | lateral occipital cortex | 126 | 5.25 | 5.1 | 48 | -62 | 34 |
| Interaction APM * ANX | positive | 122 | middle temporal gyrus | 7603 | 10.9 | Inf | -50 | -62 | 4 |
|  |  |  | anterior cingulate gyrus | 191 | 7.89 | 7.41 | 8 | 22 | 26 |
|  |  |  | superior frontal gyrus | 643 | 7.43 | 7.02 | -20 | -4 | 52 |
|  |  |  | right thalamus | 568 | 5.53 | 5.35 | -8 | -18 | 0 |
|  |  |  | left thalamus | 194 | 6.21 | 5.96 | -18 | -22 | 12 |
|  |  |  | posterior parahippocampal gyrus | 246 | 6.05 | 5.81 | 22 | -28 | -16 |
|  |  |  | superior frontal gyrus | 203 | 5.3 | 5.14 | 22 | 24 | 36 |
|  |  |  | precentral gyrus | 122 | 5.22 | 5.07 | 46 | 6 | 22 |
|  |  |  | mid insular cortex | 154 | 4.42 | 4.33 | -40 | 4 | -6 |
| Interaction APM * ANX | negative | 128 | frontal orbital cortex | 2029 | 9.69 | Inf | -44 | 36 | -10 |
|  |  |  | left hippocampus | 1219 | 9.13 | Inf | -32 | -40 | -4 |
|  |  |  | right cerebellum | 1676 | 8.15 | 7.62 | 30 | -78 | -22 |
|  |  |  | left cerebellum | 162 | 8.15 | 7.62 | -2 | -52 | -42 |
|  |  |  | orbitofrontal cortex | 1192 | 5.17 | 5.02 | 28 | 14 | -16 |
|  |  |  | right thalamus | 932 | 7.6 | 7.17 | 18 | -34 | 4 |
|  |  |  | angular gyrus | 365 | 7.02 | 6.67 | 42 | -56 | 26 |
|  |  |  | posterior cingulate | 141 | 7.01 | 6.66 | 0 | -20 | 42 |
|  |  |  | dorsal anterior cingulate gyrus | 621 | 4.97 | 4.83 | -2 | 16 | 38 |
|  |  |  | frontal pole | 286 | 6.47 | 6.19 | 34 | 34 | -8 |
|  |  |  | superior parietal lobule | 139 | 6.44 | 6.17 | -26 | -44 | 56 |
|  |  |  | left caudate | 160 | 6.29 | 6.03 | -18 | 14 | 14 |
|  |  |  | middle frontal gyrus | 303 | 6.12 | 5.88 | -44 | 28 | 28 |
|  |  |  | central operculum cortex | 179 | 5.96 | 5.74 | -42 | -14 | 16 |
|  |  |  | left cerebellum | 215 | 5.59 | 5.4 | -46 | -68 | -30 |
|  |  |  | frontal pole | 174 | 5.1 | 4.95 | 46 | 42 | 18 |
|  |  |  | middle temporal gyrus | 128 | 4.97 | 4.84 | -60 | -4 | -20 |
|  |  |  | inferior frontal gyrus (pars triangularis) | 195 | 4.82 | 4.7 | 54 | 30 | 22 |

**Supplementary Table 2 | All significant activations for clusters (P<0.001 uncorrected) with FWE cluster correction (P<0.05).** Second-level univariate analysis of autonomic perceptual mismatch for the main effect of APM and interactions of hypermobility and anxiety. Degrees of freedom 227. Co-ordinates and statistics of all clusters were generated using SPM12 in MATLAB.
